# The impact of population mobility on COVID-19 incidence and socioeconomic disparities at the sub-city level in 314 Latin American cities

**DOI:** 10.1101/2021.04.13.21255413

**Authors:** Josiah L. Kephart, Xavier Delclòs-Alió, Usama Bilal, Olga L. Sarmiento, Tonatiuh Barrientos-Gutiérrez, Manuel Ramirez-Zea, D. Alex Quistberg, Daniel A. Rodríguez, Ana V. Diez Roux

## Abstract

**Background:** Little is known about the impact of changes in mobility at the sub-city level on subsequent COVID-19 incidence or the contribution of mobility to socioeconomic disparities in COVID-19 incidence.

**Methods:** We compiled aggregated mobile phone location data, COVID-19 confirmed cases, and features of the urban and social environments to analyze linkages between population mobility, COVID-19 incidence, and educational attainment at the sub-city level among cities with >100,000 inhabitants in Argentina, Brazil, Colombia, Guatemala, and Mexico from March to August 2020. We used mixed effects negative binomial regression to examine longitudinal associations between changes in weekly mobility (lags 1-6 weeks) and subsequent COVID-19 incidence at the sub-city level, adjusting for urban environmental factors.

**Findings:** Among 1,031 sub-cities representing 314 cities in five Latin American countries, 10% higher weekly mobility was associated with 8.5% (95% CI 7.4% to 9.5%) higher weekly COVID-19 incidence the following week. This association gradually declined as the lag between mobility and COVID-19 incidence increased and was not different from the null at a six-week lag. We found evidence that suggests differences in mobility reductions are a driver of socioeconomic disparities in COVID-19 incidence.

**Interpretation:** Lower population movement within a sub-city is associated with lower risk of subsequent COVID-19 incidence among residents of that sub-city. Implementing policies that reduce population mobility at the sub-city level may be an impactful COVID-19 mitigation strategy that takes equity into consideration and reduces economic and social disruption at the city or regional level.

**Funding:** Wellcome Trust

## 1. Background

Many of the most severe COVID-19 outbreaks worldwide have occurred in the cities of North and South America,^1^ where 80% of the population lives in urban areas. In attempts to mitigate the spread of the novel coronavirus SARS-CoV-2, governments have relied heavily on regional or city-wide interventions to reduce SARS-CoV-2 transmission through universal policies such as stay-at-home orders. While these widespread measures have undoubtedly helped to mitigate COVID-19 incidence, they have also incurred enormous societal and economic costs, particularly in many cities in the Americas where rampant transmission and the associated stay-at-home orders have persisted.^2^

Managing population movement has been a central component of efforts to reduce COVID-19 transmission, based on the intuitive idea that less population mobility leads to fewer opportunities for SARS-CoV-2 transmission. Studies that have empirically quantified the impacts of mobility reductions on COVID-19 incidence have generally found positive associations between mobility and subsequent COVID-19 incidence or deaths.^3–8^ However, a majority of these studies were done at the country or provincial level, or have examined mobility within a single city.^9,10^ Understanding the relationship between mobility and COVID-19 incidence at the sub-city level is essential to evaluate the potential effectiveness of dynamic, geographically targeted policies to reduce mobility and mitigate COVID-19 incidence at a community level while minimizing regional- or city-wide disruption. To our knowledge there are no studies examining the impact of sub-national mobility on COVID-19 incidence in Latin America, and none overall that examine a large number of cities at the sub-city or neighborhood level. This research gap is particularly relevant in Latin America, where the pandemic has been persistent and the public health response to COVID-19 is shifting towards a potentially years-long struggle, even as vaccine roll-out begins.^2,11^

Cities in the Americas are home to some of the largest societal inequities in the world.^12,13^ Furthermore, significant disparities in COVID-19 incidence by socioeconomic status (SES) have emerged in cities across the world.^14^ Little is known about the contribution of inequitable mobility patterns during COVID-19 lockdowns to COVID-19 disparities by socioeconomic status.^15^ Understanding the contribution of mobility to disparities in COVID-19 incidence at the sub-city level will inform policymakers who seek actionable interventions to reduce COVID-19 disparities and promote health equity.

To address these gaps, this analysis leverages data from a rich dataset of 314 large, heterogeneous Latin American cities to 1) study the association between sub-city mobility changes during the first 6 months of the COVID-19 pandemic and subsequent COVID-19 incidence and 2) examine whether sub-city mobility changes contributed to socioeconomic disparities in COVID-19 incidence.

## 2. Methods

### 2.1. Study area

This study was conducted as part of the *Salud Urbana* en *América Latina* (SALURBAL) project. The SALURBAL project has compiled and harmonized data on health, social, and environmental characteristics for 371 cities with more than 100,000 residents in 11 Latin American countries.^16^ In this this analysis, we include 1,031 sub-city units from 314 cities in Argentina, Brazil, Colombia, Guatemala, and Mexico. Sub-city units were defined as ‘Municipios’ in Brazil, Colombia, Guatemala, and Mexico. In Argentina, sub-cities correspond to ‘Comunas’ in the Ciudad de Buenos Aires, ‘Partidos’ in the Provincia de Buenos Aires, and ‘Departamentos’ elsewhere.^17^

### 2.2. Measures and data sources

To evaluate sub-city mobility levels, we used anonymized, spatially aggregated mobile phone data provided by the United Nations Development Programme in Latin America and the Caribbean (UNDP-LAC) and Grandata.^18^ This data includes estimates of human mobility as defined by the number of out-of-home events within a given municipal area by mobile phone users on each day from March 3 - August 29, 2020, as compared to out-of-home events on a baseline, pre-COVID-19 date of March 2, 2020.^19^ Mobility within a given geographic area is defined within this data as all out-of-home events that occurred within the geographic area, regardless of whether the mobile phone user was a resident or non-resident of the geographic area.

To evaluate COVID-19 incidence at sub-city level, we used daily confirmed COVID-19 cases as reported directly by the national governments of Argentina, Brazil, Colombia, Guatemala, and Mexico. Additional information on the compilation of COVID-19 case data by the SALURBAL project and the specific data sources have been previously described.^20^

To measure socioeconomic status at the sub-city level, we used an index of educational attainment, as this reflects both differences in formality/informality in the labor market (relating to the ability to reduce mobility, such as working from home) and differences in SARS-CoV-2 exposure through increased occupational interactions with the public. The index includes an average of the z-scores of the % population aged 25 or above that has completed secondary education or above, and the % population aged 25 or above that has completed university education or above.^21^ We also examined residential overcrowding as both a metric of overall socioeconomic status as well as specifically related to risk of within-household transmission of SARS-CoV-2. We defined residential overcrowding as the % of households with more than 3 people per room. Both measures were calculated at the sub-city level using data from the latest available census. We also obtained data on the total population of the sub-city unit, obtained from population projections, and population density, defined as the number of inhabitants per square km of built-up area in the unit. Built-up area is calculated using Facebook’s Population Density Maps.^22,23^

### 2.3. Statistical Analysis

#### 2.3.1. Mobility data

Mobility estimates were originally provided as the % absolute change in out-of-home events on a given date compared to a baseline of March 2, 2020 ([(post-baseline mobility /baseline mobility) – 1] * 100). To reduce the influence of fluctuations in mobility by day of week, we calculated the weekly mean mobility within each sub-city for the week of March 2 – March 9, 2020 (i.e. pre-pandemic baseline week). For each sub-city, weekly mean mobility was calculated for each 7-day period following the date of the 2^nd^ recorded case within each sub-city (e.g. Week 1 was defined as 1 – 7 days following the date of the 2^nd^ recorded case in a sub-city, Week 2 was defined as 8-15 days following the 2^nd^ recorded case, etc.). We transformed the % absolute change in weekly mobility ([(post-baseline mobility /baseline mobility) – 1] * 100) to the log ratio of weekly vs. baseline weekly mobility (ln[(post-baseline mobility change + 1)/(baseline mobility + 1)]) during each subsequent 7-day period following the 2^nd^ recorded COVID-19 case through August 29th, 2020 (range of weeks post-baseline [1, 22]).

#### 2.3.2. COVID-19 case data

Daily COVID-19 confirmed case counts were provided by national governments. In <1% of sub-city-days, negative case values were reported, and case counts on these days were set to zero. For each sub-city, we calculated the weekly sum of confirmed cases following the date of the 2^nd^ reported case, temporally aligned with weekly mobility (Section 2.3.1).

#### 2.3.3. Analysis of mobility and COVID-19 incidence

First, we performed a longitudinal analysis of the association between weekly sub-city COVID-19 incidence (outcome) and lagged weekly sub-city mobility (primary exposure), adjusting for potential confounders at the sub-city level. We conducted a weekly time-series analysis beginning on the date of the 2^nd^ reported case in each sub-city, with mobility lagged 1 - 6 weeks behind cases in subsequent models. We adjusted for sub-city educational attainment, prevalence of residential overcrowding, population density, and country as potential confounders. We report univariate associations between each independent variable and the outcome (COVID-19 incidence) and full results from the adjusted model, with mobility lagged one week prior to cases. We also report coefficients of the adjusted association between lagged mobility and COVID-19 incidence comparing mobility lags of 1, 2, 3, 4, 5, and 6 weeks prior to COVID-19 incidence.

Second, we analyzed the contribution of mobility to the observed association between educational attainment and COVID-19 incidence. Building on the primary, adjusted model of mobility (with one-week lag) and COVID-19 incidence described above, we examined how the association between sub-city educational attainment and COVID-19 incidence changed after including mobility in the model. We subsequently stratified this analysis of SES by country to account for possible differences in national lockdown policies during the pandemic, disparities in testing coverage by socioeconomic status within and between countries, and to account for different definitions of sub-city units. We hypothesized that a negative association exists between educational attainment and COVID-19 incidence, and that adding mobility to the model would attenuate this association. If observed, this would suggest that mobility (or the inability to reduce mobility in response to lockdowns) is a contributor or partial mediator of the relationship between sub-city education and COVID-19 incidence.

In all cases, we used a mixed effects negative binomial model with random intercepts for sub-city and city, with sub-city population as an offset. All analyses were performed using R (www.r-project.org) and modeling was performed with the *glmmTMB package*.^24^

## 3. Results

### 3.1. Sub-city characteristics

The analysis included 1,031 sub-cities representing 314 cities distributed as follows: Argentina (N sub-cities = 107; N cities = 33), Brazil (416; 151), Colombia (82; 35), Guatemala (20; 3), and Mexico (406; 92). Four sub-cities (0.4 %) were excluded from the analysis due to missing data. In Table 1, we present descriptive characteristics of the sub-cities, stratifying by tertiles of cumulative COVID-19 incidence per 100k inhabitants during the study period (Mar 2 – Aug 29, 2020). During the study period, sub-cities in the analysis had a median cumulative COVID-19 incidence of 855 confirmed cases per 100k inhabitants (25^th^ percentile 401, 75^th^ percentile 1,655). The median ratio of mobility during the study period compared to baseline was 0.79 (25^th^ percentile 0.70, 75^th^ percentile 0.90) in the lowest tertile of cumulative COVID-19 incidence, 0.76 (0.65, 1.00) in the middle tertile, and 0.85 (0.65, 1.05) in the highest tertile. The median sub-city population was 130,800 inhabitants (25^th^ percentile 44,200, 75^th^ percentile 277,000) and we observed a positive association between sub-city population size and increasing cumulative COVID-19 incidence tertile. We also observed a positive association between population density and COVID-19 incidence tertile, yet a negative association between residential overcrowding and COVID-19 incidence tertile. We observed a positive association between sub-city educational attainment and COVID-19 incidence tertile, suggesting higher confirmed COVID-19 incidence among sub-cities with higher educational attainment.

**Table 1.** Sub-city characteristics, overall and stratified by cumulative COVID-19 incidence (cases per 100k inh.) between March 2^nd^ and August 29^th^, 2020 among 1,031 sub-cities in 314 Latin American cities

|  | Total | Cumulative COVID-19 incidence (cases per 100k inhabitants) by tertiles |  |  |
| --- | --- | --- | --- | --- |
|  |  | Lowest (0, 506.6] | Middle (506.6, 1,314.1] | Highest (1,314.1, 9,403.5] |
| Sub-cities (n) | 1,031 | 344 | 343 | 344 |
| Cumulative COVID-19 cases per 100k inh.<sup>a, b</sup> | 855 [401, 1,655] | 307 [183, 401] | 855 [664, 1,081] | 2,023 [1,656, 2,602] |
| Change in mobility (ratio)<sup>a, c</sup> | 0.80 [0.67, 0.99] | 0.79 [0.70, 0.90] | 0.76 [0.65, 1.00] | 0.85 [0.65, 1.05] |
| Population (thousands)<sup>a</sup> | 130.8 [44.2, 277.0] | 83.6 [28.4, 181.5] | 136.3 [46.3, 304.1] | 176.6 [82.7, 345.4] |
| Pop. density (pop. in thousands / sq.km)<sup>a</sup> | 8.2 [5.5, 14.0] | 7.3 [5.5, 11.4] | 9.0 [5.8, 15.5] | 8.1 [5.2, 15.3] |
| Residential overcrowding (%)<sup>a, d</sup> | 5.3 [2.8, 10.9] | 10.9 [6.7, 14.7] | 5.2 [2.8, 9.8] | 3.1 [1.7, 5.0] |
| Population educational attainment<sup>a, e</sup> | -0.84 [-1.60, -0.02] | -1.38 [-2.12, -0.56] | -0.66 [-1.35, 0.14] | -0.56 [-1.18, 0.26] |

<sup>a</sup> Values correspond to medians [Q1, Q3].
<sup>b</sup> Between March 2<sup>nd</sup> and August 29<sup>th</sup>, 2020.
<sup>c</sup> Ratio of overall vs baseline mobility (March 2<sup>nd</sup>-9<sup>th</sup>, 2020).
<sup>d</sup> Proportion of households with more than 3 people per room.
<sup>e</sup> Index that includes an average of the z-scores of the % population in the sub-city aged 25 or above that has completed secondary education or above, and the % population aged 25 or above that has completed university education or above.

### 3.2. Sub-city mobility levels and COVID-19 incidence

Descriptive plots of sub-city changes in mobility and of COVID-19 incidence are presented in Figure 1, stratified by country. As referenced on the Y-axis, the median sub-cities within all countries experienced a substantial decrease in mobility during the end of March, as news of the pandemic spread quickly across the world and lockdown policies were implemented in many settings. However, there is wide variation between countries and between sub-cities within countries regarding the duration and patterns of the mobility reductions, represented in Figure 1 by the upper (75^th^ percentile) and lower (25^th^ percentile) boundaries of color shading. Similarly, referenced on the X-axis is the 7-day moving average confirmed daily COVID-19 cases per 100k inhabitants. There is substantial variation in patterns of incidence between countries, with incidence declining by the end of the study period in some countries but continuing to rise in others. There is also wide variation in incidence rates among sub-cities in the same country, similarly represented in Figure 1 by the upper (75^th^ percentile) and lower (25^th^ percentile) boundaries of color shading.

**Figure 1.**
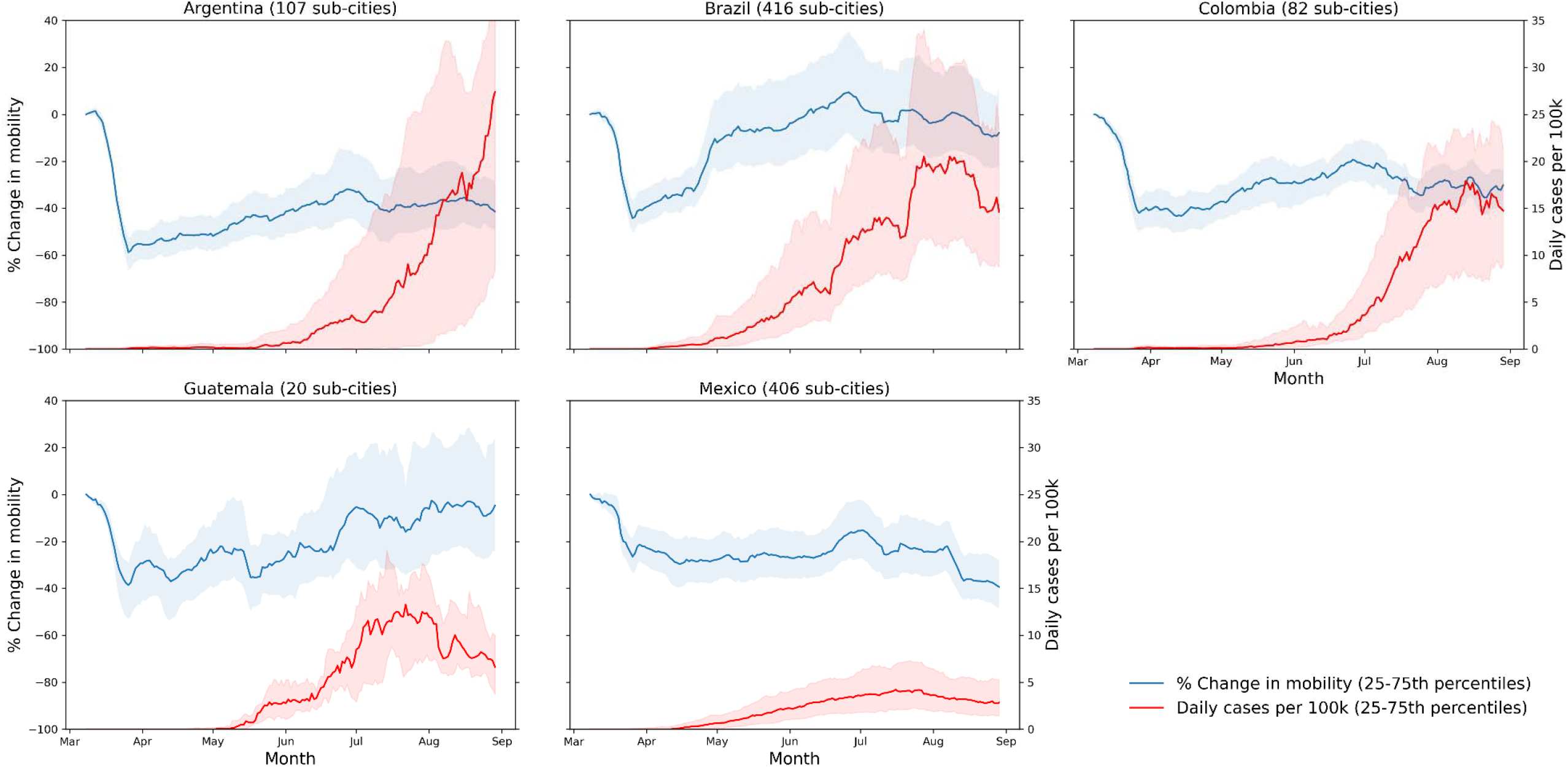
Daily change in mobility from baseline date and daily cases per 100k inhabitants among 1,031 sub-cities in 314 cities in five Latin American countries, 7-day moving averages between March 2^nd^ and August 29^th^, 2020.

The univariate associations between independent variables and confirmed COVID-19 cases as well as the results of our adjusted model of the association between mobility (lagged one week) and subsequent COVID-19 incidence are presented in Table 2. Coefficients from the negative binomial model have been exponentiated and are interpretable as Incidence Rate Ratios (IRR). In the adjusted model, we observed an IRR of 2.35 (95% CI 2.12 to 2.60) COVID-19 incidence per log unit increase in mobility during the prior week. This implies that a 10% higher mobility is associated with 8.5% (95% CI 7.4% to 9.5%) higher COVID-19 incidence in the subsequent week (1.10 ^ [ln(2.35)] = 0.085). Notably, we observed no associations between COVID-19 incidence and population density (IRR 1.00 (95% CI 0.99 to 1.01)) or residential overcrowding (IRR 0.99 (95% CI 0.98 to 1.00)) in the adjusted model. We observed a positive association between sub-city educational attainment and COVID-19 incidence (IRR 1.07 (95% CI 1.03 to 1.11)), suggesting higher confirmed COVID-19 incidence in sub-cities with higher education. In the analysis comparing the effects of mobility with lags of 1 - 6 weeks prior to incidence, the association between lagged mobility and subsequent COVID-19 incidence was strongest with a lag of one week (Figure 2) in the adjusted models. This association gradually decayed through weeks two to five, until a lag of six weeks for which we observed no association between mobility and COVID-19 incidence.

**Table 2.** Incidence Rate Ratios of unadjusted and adjusted associations between sub-city COVID-19 weekly incidence (outcome), weekly mobility (primary exposure), and sub-city characteristics from mixed effects negative binomial models.

|  | Unadjusted <sup>a</sup> |  | Adjusted <sup>b</sup> |  |
| --- | --- | --- | --- | --- |
|  | IRR | 95% CI | IRR | 95% CI |
| Mobility change from baseline (1-week lag) (log of ratio) | 8.12 | (6.95, 9.49) | 2.35 | (2.12, 2.60) |
| Weeks since 2<sup>nd</sup> case | 1.18 | (1.17, 1.18) | 1.17 | (1.16, 1.17) |
| Population density (pop. in thousands / sq. km) | 1.01 | (1.00, 1.01) | 1.00 | (0.99, 1.01) |
| Residential overcrowding (%) | 0.96 | (0.95, 0.96) | 0.99 | (0.98, 1.00) |
| Population educational attainment (z-score) | 1.12 | (1.10, 1.15) | 1.07 | (1.03, 1.11) |

<sup>a</sup> Unadjusted models include sub-city weekly COVID-19 incidence (outcome) and each single variable (exposure) for 1,031 sub-cities in Latin America, using longitudinal mixed effects negative binomial models with random intercepts for sub-city, with sub-city population as an offset.
<sup>b</sup> Adjusted models include sub-city weekly COVID-19 incidence (outcome), mobility (primary exposure), weeks since 2<sup>nd</sup> case, population density, residential overcrowding, population educational attainment, and country for 1,031 sub-cities in Latin America, using longitudinal mixed effects negative binomial models with random intercepts for sub-city and city, with sub-city population as an offset.

**Figure 2.**
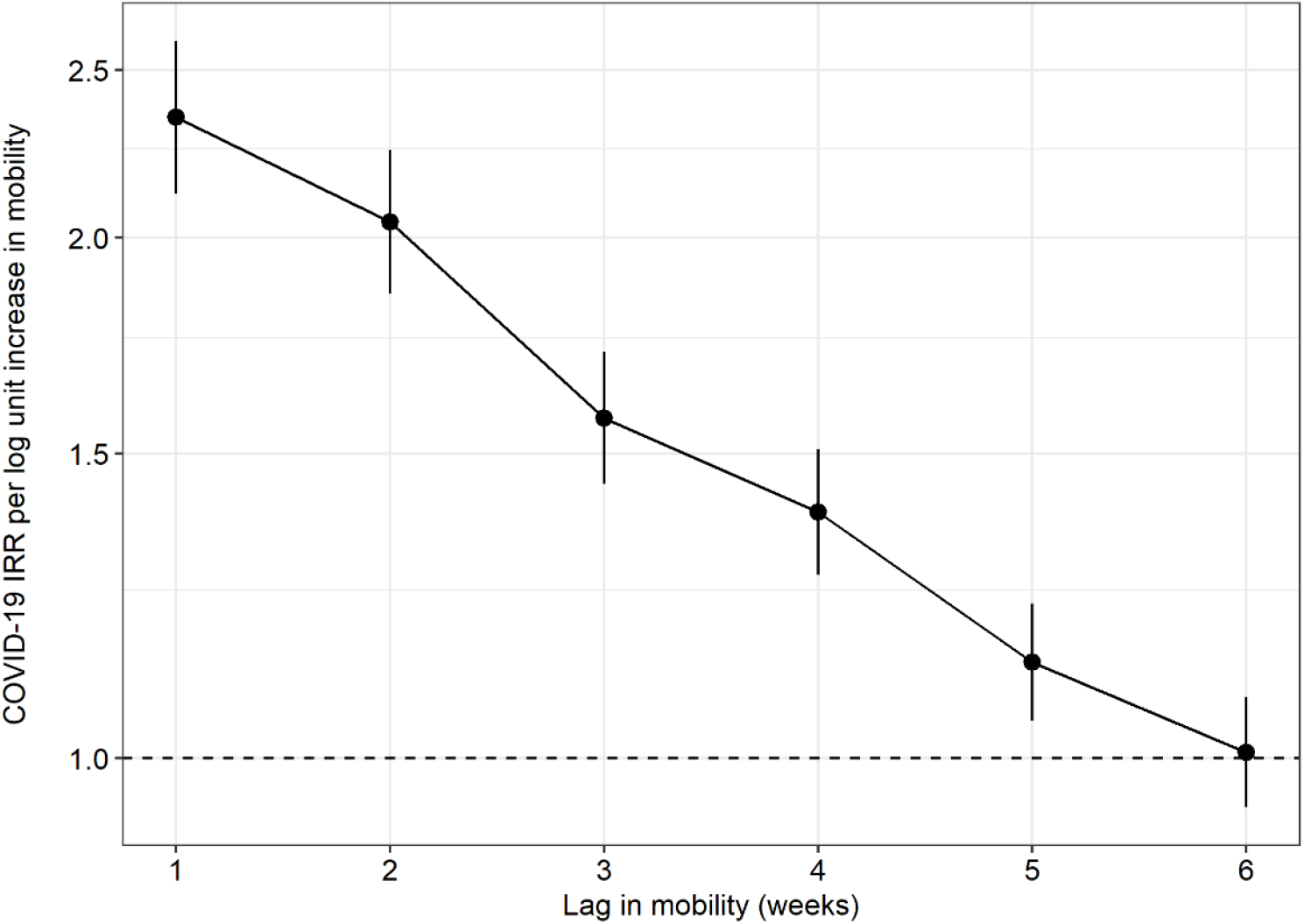
Incidence rate ratios (IRR) and 95% confidence intervals of the adjusted association between lagged mobility change and COVID-19 incidence at the sub-city level, comparing mobility from one to six weeks prior to COVID-19 incidence^a^. ^a^Results come from adjusted models that include sub-city weekly COVID-19 incidence (outcome), mobility (primary exposure), weeks since 2^nd^ case, population density, residential overcrowding, educational attainment, and country for 1,031 sub-cities in Latin America, using longitudinal mixed effects negative binomial models with random intercepts for sub-city and city, with sub-city population as an offset. Weekly mobility was lagged from one to six weeks prior to weekly COVID-19 incidence in subsequent models.

### 3.3. Contribution of sub-city mobility changes to socioeconomic disparities in COVID-19 incidence

The associations between sub-city educational attainment and COVID-19 incidence are presented overall for all sub-cities and stratified by country in Figure 3. Without adjusting for mobility in the models (red interval estimates), we observed a negative association between sub-city educational attainment and COVID-19 incidence (IRR < 1, i.e. higher education, lower incidence) overall. After adjusting for mobility (blue interval estimates), this negative association between sub-city education and COVID-19 incidence was qualitatively changed to a positive association, indicating that differences in mobility contribute to the association between sub-city education and COVID-19 incidence overall. The association between sub-city education and COVID-19 incidence varied widely between countries. In Brazil and Colombia, we observed a negative association between sub-city education and COVID-19 incidence which was attenuated upon adjusting for mobility in Brazil but less so in Colombia. In Mexico, the positive association between sub-city education and COVID-19 incidence without mobility was strengthened with the adjustment for mobility.

**Figure 3.**
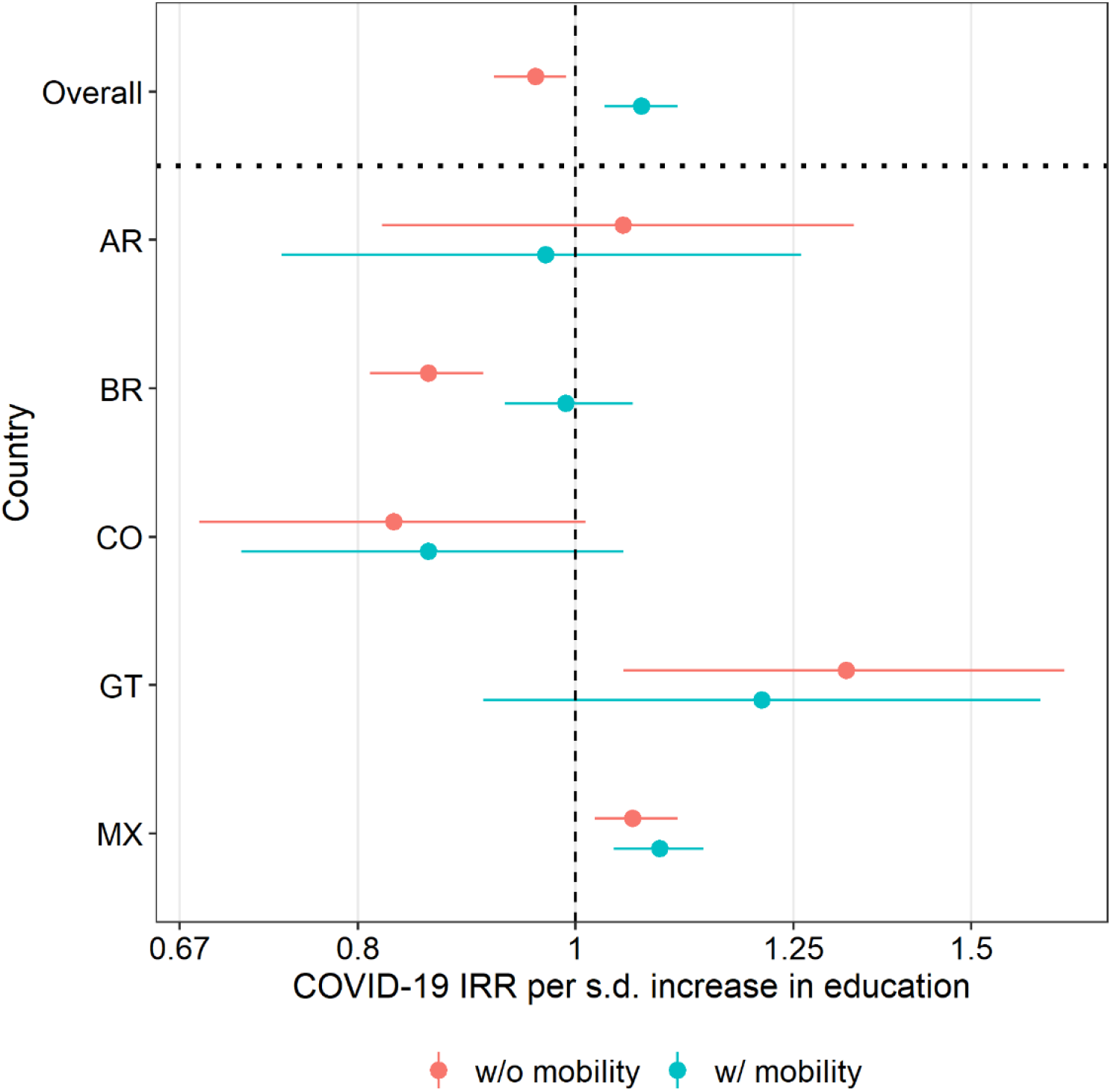
COVID-19 Incidence Rate Ratio with 95% confidence intervals per s.d. higher education attainment at the sub-city level, stratified by country and comparing non-adjustment versus adjustment for mobility one week prior to COVID-19 incidence ^a^. ^a^Results come from adjusted models that include sub-city weekly COVID-19 incidence (outcome), mobility lagged one week prior to COVID-19 incidence, weeks since 2^nd^ case, population density, residential overcrowding, educational attainment, and country for 1,031 sub-cities in Latin America, using longitudinal mixed effects negative binomial models with random intercepts for sub-city and city, with sub-city population as an offset. The adjusted COVID-19 incidence rate ratio (IRR) of a one s.d. increase in sub-city education attainment are presented overall and stratified by country, with and without adjustment for sub-city mobility.

## 4. Discussion

In this study, we examined the impact of human mobility at the sub-city level on subsequent COVID-19 incidence in a multicity, multicountry analysis. We observed a strong, positive association between population mobility within a sub-city and subsequent COVID-19 incidence among residents of that sub-city. This association was strongest with a mobility lag of one week, where a 10% higher mobility was associated with an 8.5% (95% CI 7.4% to 9.5%) higher COVID-19 incidence. The IRR of increased mobility on COVID-19 incidence gradually decayed across subsequent weekly lags until six weeks. These results provide evidence that mobility is a contributor to COVID-19 incidence at the sub-city level, and that the sub-city level may be an effective subject for targeted interventions which reduce mobility to mitigate SARS-CoV-2 transmission, thereby potentially limiting city-wide disruption. We also observed evidence that suggests differences in the ability (or lack of ability) to reduce sub-city level mobility by sub-city level educational attainment may contribute to COVID-19 disparities overall, though this association varied by country. Interventions that reduce disparities in mobility may serve as effective strategies to reduce subsequent disparities in COVID-19 incidence.

We observed an association between weekly mobility and weekly COVID-19 incidence that was strongest with a mobility lag of one week and gradually decayed through a lag of six weeks. This is similar to a county-level analysis in the USA which found high correlations (> 0.7) between COVID-19 growth rate and a mobility lag of only four days prior, with the strongest correlations at mobility lags of 9-12 days.^3^ A provincial-level analysis in Italy found that highly affected provinces recorded decreases in COVID-19 incidence 9-10 days after mobility reductions from lockdown policies.^25^ These ranges are consistent with the reported COVID-19 latency period of a median of 5.1 days from infection to development of symptoms.^26^ Beyond symptom development, we would expect the average time from infection to confirmation of a COVID-19 case to vary depending on COVID-19 testing access and speed.

Reports of confirmed COVID-19 cases were gathered directly from official governmental sources, however determining the true incidence of COVID-19 in these cities is challenging for a variety of reasons, including the potential lack of access to testing and healthcare or the prevalence of asymptomatic individuals who may have not sought medical treatment. These disparities in actual versus confirmed cases are likely to be unequally distributed between and within countries and cities in this study. It would also be expected that in many settings lower socioeconomic status may increase barriers to adequate COVID-19 testing.^27,28^ We suspect that the positive association observed in the country-pooled analysis between educational attainment and COVID-19 incidence after adjusting for mobility may be partially driven by increased testing coverage in sub-cities with higher education attainment. When stratifying by country, a positive association between SES and COVID-19 incidence is strongest in Mexico, which may be driving the overall association. If cases are in fact underreported in sub-cities with lower SES, this would result in a biased estimate of the association between education and COVID-19 incidence.

The underreporting of cases linked to limited testing or differential access to testing by education could have affected not only the educational gradients in COVID-19 incidence that we observed but also the contribution of differences in mobility to those educational gradients. The fact that the negative association with education became positive after adjustment for mobility suggests that higher mobility (or smaller reductions in mobility) in low education areas is contributing to differences by education in COVID-19 incidence. Underreporting could also have impacted our estimates of the associations of mobility with COVID-incidence, if mobility is patterned by SES and SES is linked to underreporting of cases. Furthermore, our estimates of mobility do not distinguish by the mode of transportation used. Since public transportation use is more common in lower SES areas, these differences in exposure may also help explain variation in COVID-19 incidence by SES. Given disparities in COVID-19 testing access by socioeconomic status, the results we present of the magnitude of the overall mobility-incidence relationship are likely conservative estimates.

This study is strengthened by its use of a rich dataset spanning 1,031 sub-cities in 314 cities across five countries that have experienced some of the most severe COVID-19 outbreaks globally. This study also uses daily mobility data compiled from anonymized mobile phones, which directly measures community-level behavior that contributes to SARS-CoV-2 transmission. Linking these datasets allows us to conduct longitudinal analyses to provide needed empirical evidence on the effectiveness of a widespread COVID-19 mitigation strategy at a relatively fine spatial level.

This study has several limitations. Because of the lack of individual-level data, we were unable to examine the association between individual mobility, education, and COVID-19 incidence. We are limited to contextual factors reflecting the sociodemographic composition of sub-cities. Additionally, Grandata mobility datasets are metrics of the total number of out-of-home events that occur in a particular sub-city unit at a particular moment in time, regardless of travelers’ sub-city unit of residence. This means that we were unable to measure the mobility levels of residents of a particular sub-city unit, but instead we measured mobility that occurred by anyone in a particular sub-city at a given moment in time. However, this sub-city level analysis remains highly relevant to policymakers, who may be similarly focused on regulating all mobility within certain geographic areas versus regulating mobility based on an individual’s location of residence. Furthermore, because the mobility data is available at the sub-city level only, we were unable to examine mobility at a city level to evaluate the relative importance of city-level versus sub-city level mobility reductions on COVID-19 incidence. Finally, while other non-pharmaceutical interventions (NPIs), such as face-coverings, social distancing, and handwashing, play substantial roles in mitigating community-level COVID-19 incidence, these behaviors are challenging to measure.^29^ We expect that mobility and other NPIs are correlated in timing and adherence and have distinct impacts on COVID-19 incidence, which are difficult to disentangle.^8,30^ However, policies that directly intervene on mobility are core actionable interventions and, in many situations, may be a first line of defense at a city or sub-city level.

## 5. Conclusion

In this study of more than 1,000 sub-cities across 314 cities of five Latin American countries, we observed a positive association between weekly mobility changes and COVID-19 incidence. Specifically, a 10% higher weekly mobility was associated with an 8.5% higher COVID-19 incidence the following week, and this association decayed with longer lags. Furthermore, we found evidence that suggests differences in mobility contribute to COVID-19 incidence disparities by educational attainment at the sub-city level, but this requires further confirmation in settings with higher testing accessibility. This analysis provides novel evidence that across a wide range of cities diverse in health infrastructure, COVID-19 progression, and government response to the pandemic, out-of-home population movement within sub-cities increases the risk of COVID-19 incidence to residents of those sub-cities.

Policies which aim to reduce SARS-CoV-2 transmission and mitigate COVID-19 disparities may be effective at the sub-city or neighborhood level, which may help limit regional or city-wide disruption. Furthermore, reducing sub-city sociodemographic disparities in the ability to reduce mobility, such as economic support that facilitates the ability of individuals to stay at home, may serve as effective strategies to reduce subsequent disparities in COVID-19 incidence. Further research is warranted to explore the effectiveness of dynamic, geographically targeted policies to reduce SARS-CoV-2 transmission and disparities on a population level.

## Data Availability

Mortality data are available upon request.

## Contributors

JK, XDA, UB, and ADR conceptualized the analysis. JK and XDA had access to all data, verified the data, completed the formal analysis, and created the original draft the manuscript. ADR and DR acquired funding and provided supervision. All authors contributed to data gathering, methodology, and manuscript editing.

## Acknowledgements

The authors acknowledge and thank the United Nations Development Programme – Latin America and the Caribbean (UNDP-LAC) and GRANDATA for providing the mobility datasets used in this analysis. Furthermore, the authors acknowledge the contribution of all SALURBAL project team members. For more information on SALURBAL and to see a full list of investigators, see https://drexel.edu/lac/salurbal/team/. SALURBAL acknowledges the contributions of many different agencies in generating, processing, facilitating access to data or assisting with other aspects of the project. Please visit https://drexel.edu/lac/data-evidence for a complete list of data sources. The SALURBAL/Urban Health in Latin America project is funded by the Wellcome Trust (205177/Z/16/Z). U.B. was also supported by the Office of the Director of the National Institutes of Health under award number DP5OD26429. The funding sources had no role in the analysis, writing or decision to submit the manuscript.

